# Should a repeat cervical cerclage be inserted when the primary cerclage fails, to prevent pregnancy loss and preterm birth? A systematic review and meta-analysis

**DOI:** 10.1101/2025.08.05.25332738

**Authors:** A Emms, M Vaughan, R Man, RK Morris, V Hodgetts-Morton, N Pilarksi

**Affiliations:** Women’s Division, Birmingham Women’s and Children’s NHS Foundation Trust, Birmingham UK; Department of Applied Health Sciences, School of Health Sciences, College of Medicine and Health, University of Birmingham, UK; University of Birmingham Medical School, University of Birmingham, UK

## Abstract

**Introduction:** Spontaneous preterm birth occurs in 0.5-1% of pregnant women and is commonly attributed to cervical insufficiency. Cervical cerclage can reduce the rate of spontaneous preterm birth in high-risk women with a shortened cervix. Management options when primary cerclage fails are uncertain. This review aims to synthesise the evidence for repeat cervical cerclage to understand outcomes and aid decision making for patients and clinicians.

**Materials and Methods:** Databases were searched according to a prospective protocol registered with PROSPERO (CRD42025638147). Included studies reported outcomes for pregnant women with a cervical cerclage in situ that failed and compared repeat cervical cerclage with expectant management. The primary outcome was a composite outcome of pregnancy loss; to include miscarriage, stillbirth, neonatal death and termination of pregnancy. Secondary outcomes included preterm birth less than 37 and less than 34 weeks, miscarriage and previable neonatal death less than 24 weeks, birthweight and gestational age at delivery. Random effects meta-analysis was performed using RevMan and risk of bias was assessed using the Robins-I tool.

**Results:** Database and citation searching retrieved 1006 titles and abstracts. There were 20 papers that underwent full text review. Six retrospective cohort studies met inclusion criteria for meta-analysis. There was no significant difference in pregnancy loss (OR 1.65 95% CI 0.23-11.62), preterm birth less than 34 weeks (OR 1.11 95% CI 0.14-8.70) or preterm birth less than 37 weeks(OR 1.88 95% CI 0.74-4.80) for repeat cervical cerclage compared to expectant management, with a trend towards improved outcomes with expectant management.

**Conclusions:** There was no evidence of any difference in pregnancy loss or preterm birth with or without repeat cervical cerclage. The overall quality and quantity of evidence is poor and patients should be informed of this. Further research in this area is required for informed decision making.

## 1. Introduction

Prematurity is the leading cause of child mortality and morbidity worldwide, and accounts for approximately 10% of all births. (1) The aetiology of preterm birth (PTB) is multifactorial; however premature dilatation of the cervix, described as cervical insufficiency, weakness or incompetence, is commonly attributed as the primary causative factor in spontaneous preterm birth (sPTB). This occurs in 0.5-1% of pregnant women. (2)

For decades, insertion of a cervical cerclage (either low vaginal, high vaginal or transabdominal) has been used as a management strategy to prevent this process. Cervical cerclage has been shown to reduce the rate of sPTB in women at high risk of preterm birth with a shortened cervix demonstrated on ultrasound. (2-4) It is, however, uncertain regarding what the next step should be when the first cerclage fails in women who show no signs or symptoms of threatened preterm labour; should a repeat cerclage be performed if the woman is eligible for one, or not?

There is no consensus nationally in the UK to define primary cerclage failure: either by transvaginal cervical length ultrasound surveillance or by physical speculum examination of the cervix. Once failure is diagnosed there is no guidance for the most appropriate management strategy for prevention of pregnancy loss and preterm birth. (5, 6) Clarfield et al have published the largest study to date comparing repeat cervical cerclage with expectant management. (7) This study included 40 women who had reinforcing cerclage after ultrasound evidence of failure and 40 women who received expectant management. Their data showed no significant differences in pre-viable or preterm delivery, however those who had repeat cerclage had increased rates of placental infection and chorioamnionitis.

This review aims to synthesise the current body of evidence investigating outcomes of repeat cervical cerclage compared with expectant management, following asymptomatic primary cerclage failure. We aim ultimately to be able to draw conclusions to guide clinical decision making when considering a repeat cerclage or not.

## 2. Methods

### 2.1 Study design and registration

This is a systematic review and meta-analysis comparing the pregnancy loss and preterm birth rates of repeat cervical cerclage with expectant management in women with asymptomatic failure of a primary cervical cerclage. It was conducted in accordance with the ‘Preferred Reporting Items for Systematic reviews and Meta-Analyses’ (PRISMA) guidelines 2009. (8, 9) Ethical approval was not required. It was prospectively registered with the International prospective register of systematic reviews (PROSPERO) prior to full data extraction and analyses. PROSPERO registration number: CRD42025638147. (10)

### 2.2 Inclusion and exclusion criteria

Given the rare incidence of this condition, and even rarer clinical scenario of considering a repeat cervical cerclage when the primary cerclage fails, all types of studies were included in the review. These included: randomised controlled trials, quasi-randomised controlled trials, cohort studies, case-control studies and case series where five or more cases were reported. Systematic reviews were excluded. Papers reporting outcomes for a cohort of patients who received repeat cervical cerclage, but did not have a comparator group of patients with primary cerclage failure who received expectant management were excluded from the meta-analysis, however were included in the qualitative overview. There were no non-English language studies eligible for inclusion.

### 2.3 Outcomes

The primary outcome was pregnancy loss. This is a composite outcome that encompasses: miscarriage or previable neonatal death (before 24 weeks gestation), stillbirth (intrauterine death after 24 weeks gestation), termination of pregnancy (medically indicated termination due to prematurity and its complications, or consequence of cerclage insertion) and early neonatal death (up to seven days post-delivery). This was chosen as our primary outcome because of the importance for patients and clinical significance.

Our pre-defined secondary outcomes encompassed a wide range of both maternal and neonatal outcomes, including those detailed in the core outcome set for interventions to prevent preterm birth, published by van Hooft et al. (11) These are listed in full in Appendix 1. Included studies reported: gestational age at delivery including less than 28 weeks, less than 32 weeks, less than 34 weeks and less than 37 weeks in live births more than 24 weeks gestation, antenatal steroid use, preterm pre-labour rupture of membranes (PPROM) and chorioamnionitis, early and late neonatal death, birthweight in grams, neonatal unit admission following live birth and neonatal unit length of stay and whether respiratory support was required.

### 2.4 Search strategy

A systematic search of the four main databases, MEDLINE, Embase, CINAHL and Cochrane, from inceptionto January 2025 was undertaken. Keywords and variants of “pregnancy”, “cervical cerclage” and “reoperation” were used, including exploded medical subject headings (MeSH), in addition to searches within the paper title, abstract and keyword fields. An example of the full search strategy is detailed in Appendix 2, which was adapted to each database searched. There were no restrictions on language, country of origin, publication date, study design or follow-up period.

### 2.5 Study selection and data extraction

Two reviewers independently reviewed the titles and abstracts in the retrieved papers (AE and MV) and any conflicts were resolved by a third independent reviewer (NP). This process was repeated for full text screening and data extraction. Covidence software was used for study screening. Data from the studies that met inclusion criteria were extracted using a piloted data entry form and then transferred onto an electronic spreadsheet.

### 2.6 Risk of bias assessment

Two review authors independently assessed risk of bias for each included study (AE and MV) with conflicts resolved by a third reviewer (NP). Risk of bias was assessed using the ROBINS-I tool for non-randomised studies, reported at individual domain level. (12)

### 2.7 GRADE certainty of evidence assessment

Certainty of evidence was assessed using the Grading of Recommendations Assessment, Development and Evaluation tool (GRADE). (13, 14)

### 2.8 Publication bias

Publication bias was not formally assessed due to there being only six studies included in the meta-analysis.

### 2.9 Data analysis

Data was synthesised using Cochrane Review Manager software (RevMan Web version 9.3.0). For assessment of dichotomous outcomes, odds ratios (ORs) were calculated, with 95% confidence intervals. Means and standard deviations were extracted for continuous outcomes. Where continuous outcomes were expressed as the median and interquartile range, these were converted to mean and standard deviation estimations according to Wan et al’s method. (15) A random effects model meta-analysis was performed, using the Mantel-Haenszel method for dichotomous outcomes, and the DerSimonian and Laird method for continuous outcomes. Heterogeneity in the estimates of effect was assessed using I^2^.

## 3. Results

### 3.1 Included studies

Following database searches, 1006 titles and abstracts were screened. Twenty papers were retrieved for full text review. Six retrospective cohort studies were eligible for inclusion in the meta-analysis. (7, 16-20) including 238 participants. Figure 1 demonstrates the search and selection process generated using Covidence software (aligned to PRISMA 2020).

**Figure 1:** Study flow diagram.

Characteristics of included studies are demonstrated in Table 2. Two studies were conducted in the United States of America (USA) (Baxter et al, Contag et al), one in Canada (Clarfield et al), one in the United Kingdom (UK) (Simcox et al), one in Korea (Song et al), one in China (Tang et al). Three studies defined primary cerclage failure as a short cervix less than 25mm in length (Baxter, Clarfield and Contag et al) (7, 16, 17)) and three defined failure as prolapse of fetal membranes through the level of the cerclage, either diagnosed by transvaginal ultrasound or by physical examination (Simcox, Song and Tang et al (18-20)). Studies used varying combinations of indications for primary cervical cerclage: either history, ultrasound-indicated or emergency cerclage and included various surgical techniques: low vaginal, high vaginal or transabdominal. One study, Song et al, included one set of twins in each study group. (19) All of Tang et al’s participants were twin pregnancies. (20) The remainder of the studies excluded multiple pregnancies. There was limited and variable reporting of baseline characteristics between the studies (supplementary file 1).

**Table 2:** Characteristics of included studies.

| Study Year (Country) | Study Design | Definition of cerclage failure | Population (Singleton/multiple pregnancies, total participants, number in each study group, total births, total live births) | Outcomes reported | Inclusion criteria | Exclusion criteria | Indication for primary cerclage |
| --- | --- | --- | --- | --- | --- | --- | --- |
| Baxter 2005 (USA) | Retrospective cohort study<br>Funding: none reported | Short cervical length <25mm on TVUS | Singleton<br>Participants: 24<br>Intervention: 5<br>Comparator: 19<br>Total births: 24<br>Total live births: 17 | Pregnancy loss, preterm birth <35 weeks | Short cervical length <25mm TVUS, GA <24 weeks | Initial cerclage US-indicated or emergency, multiple pregnancy, concomitant pessary use, incomplete records | History (inserted at 11-15 weeks based on previous history of mid-trimester loss/preterm birth <35 weeks/previous cerclage/previous failed cerclage |
| Clarfield 2024 (Canada) | Retrospective cohort study<br>Funding: none reported | Short cervical length <25mm before 24 weeks gestation | Singleton<br>Participants: 80<br>Intervention: 40<br>Comparator: 40<br>Total births: 80<br>Total live births: not possible to determine from the data | Pregnancy loss, preterm birth <37 weeks | All cases with 2 or more cerclage procedures in one pregnancy | Multiple pregnancy, if received more than 2 cerclage procedures in the study period, TAC or Shirodkar cerclage, congenital abnormalities, cerclage procedures where 2 or more stitches placed at the same time, any delivery that occurred outside the hospital | Not identified |
| Contag 2015 (USA) | Retrospective cohort study<br>Funding: none reported | Short cervical length <25mm on TVUS | Singleton<br>Participants: 51<br>Intervention: 20<br>Comparator: 31<br>Total births: 51<br>Total live births: 51 | Preterm birth <34 and <37 weeks | Singleton pregnancy, no congenital abnormalities, between 14-24 weeks gestation | Multiple pregnancy, preterm labour, rupture of membranes, intrauterine infection, congenital uterine or fetal anomalies, indication for cerclage unclear, delivery outside of base hospital, debris noted in the amniotic fluid on ultrasound | History, ultrasound or physical examination-indicated |
| Simcox 2012 (UK) | Retrospective cohort study<br>Funding: none reported | Membranes prolapsing beyond the level of | Singleton<br>Participants: 25<br>Intervention: 13 | Pregnancy loss, preterm birth <28, | Singleton pregnancy, US evidence of membranes prolapsing beyond a | Multiple pregnancy, incomplete records, those who did not fit criteria | History, ultrasound or transabdominal cerclage |
|  |  | cervical cerclage | Comparator: 12<br>Total births: 25<br>Total live births: 17 | <32, <34<br>and <37<br>weeks | cervical cerclage |  |  |
| Song<br>2010<br>(Korea) | Retrospective cohort study<br>Funding: none reported | Membranes prolapsing through the cervix visible on speculum | Both singleton and multiple (1 set of twins in each group)<br>Participants: 22<br>Intervention: 11<br>Comparator: 11<br>Total births: 24<br>Total live births: 14 | Pregnancy loss | Gestational age 16-26 weeks, cervical dilatation >1cm with bulging membranes, intact membranes, no uterine contractions, no vaginal bleeding, no fetal anomalies | Chorioamnionitis, preterm prelabour rupture of membranes, persistent contractions, GA >27 weeks | Ultrasound or physical examination-indicated |
| Tang<br>2024<br>(China) | Retrospective cohort study<br>Funding: National Natural Science Foundation of China, Shanghai Municipal Science and Technology Commission Research Fund, Program of Shanghai Academic/Technology Research Leader, Key Research Project of Pudong New Area Population and Family Planning Commission | Dilated external os to 4cm or less during follow-up | Multiple (all twin pregnancies)<br>Participants: 36<br>Intervention: 12<br>Comparator: 24<br>Total births: 72<br>Total live births: 72 | Preterm birth <34 and <37 weeks | External os dilated to 4cm or less after the initial US-indicated cerclage without signs of labour, placental abruption or chorioamnionitis, before 28 weeks gestation | Signs of labour, placental abruption or chorioamnionitis | Ultrasound (Cervical length 15mm or less) |

Table abbreviations: TVUS = transvaginal ultrasound, GA = gestational age, US = ultrasound, TAC = transabdominal cerclage, PPROM = preterm prelabour rupture of membranes.

### 3.2 Risk of bias

Risk of bias of the included studies was assessed using the ROBINS-I tool for bias assessment of observational studies. Appendix 3 demonstrates the quality of each study according to individual ROBINS-I domain assessment. None of the studies were deemed at low risk of bias for confounding. Overall, two studies were deemed at moderate risk of bias (Contag, Tang (17, 20)), two studies at serious risk of bias (Baxter, Song (16, 19)) and two studies at critical risk of bias (Clarfield, Simcox (7, 18)). Due to the limited available evidence, all six studies were included in the meta-analysis including those deemed to be at high risk of bias.

### 3.3 GRADE certainty of evidence assessment

The level of certainty of evidence was assessed using the GRADE assessment tool and applied to outcomes with three or more studies included in the meta-analysis: pregnancy loss as composite, preterm birth less than 37 weeks gestation, preterm birth less than 34 weeks gestation, miscarriage and previable neonatal death less than 24 weeks gestation, birthweight and gestational age at delivery. All outcomes were graded at very low certainty of evidence, except from miscarriage and previable neonatal death less than 24 weeks which was graded at low certainty. The full GRADE assessments with explanations for the grading are displayed in Table 3. The studies were deemed at either moderate, serious or critical risk of bias, and there was considerable heterogeneity in the estimates of effect with all outcomes except miscarriage and previable neonatal death <24 weeks. The variation between studies with their design and definitions also contributed to downgrading for indirectness.

**Table 3:**
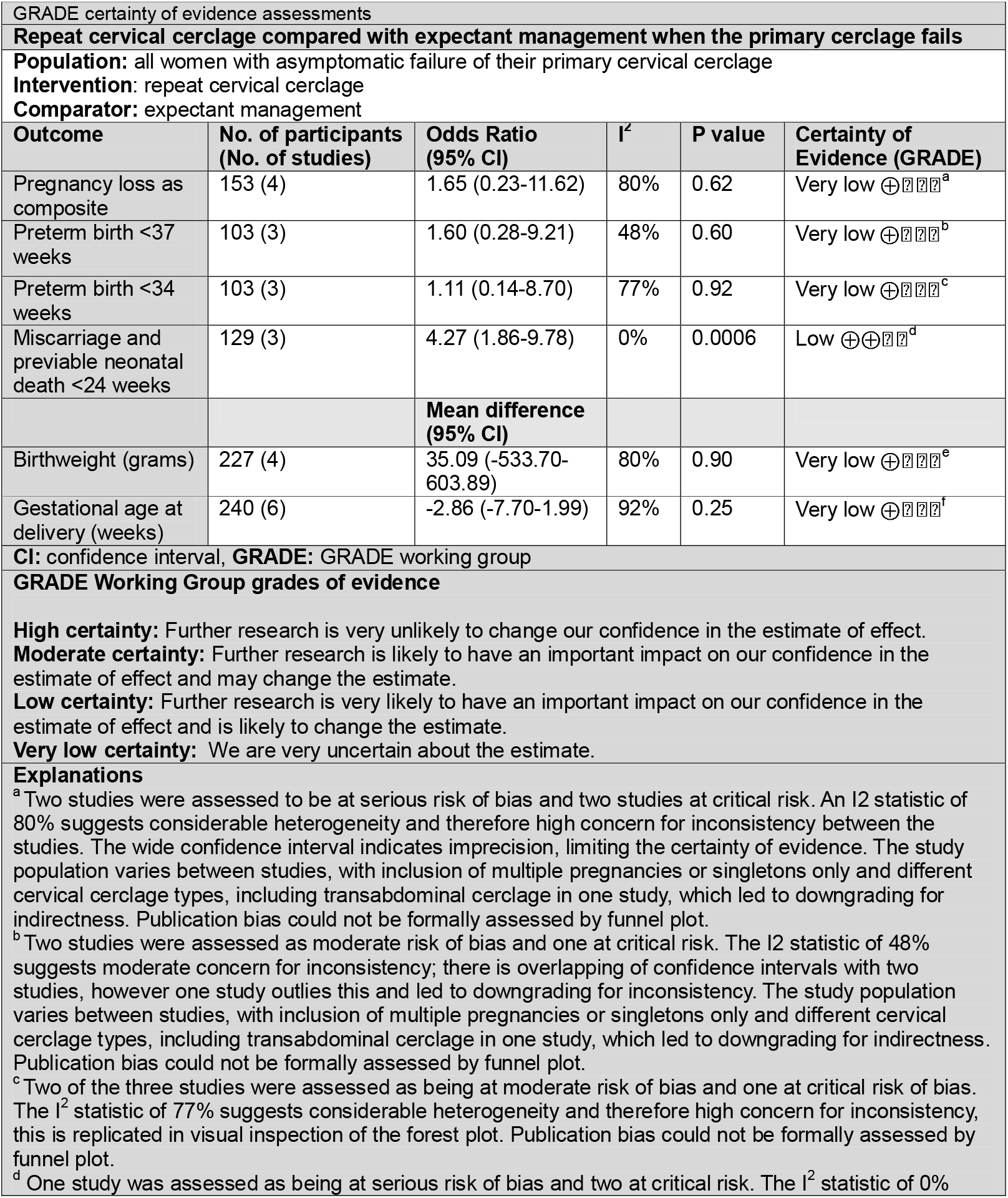
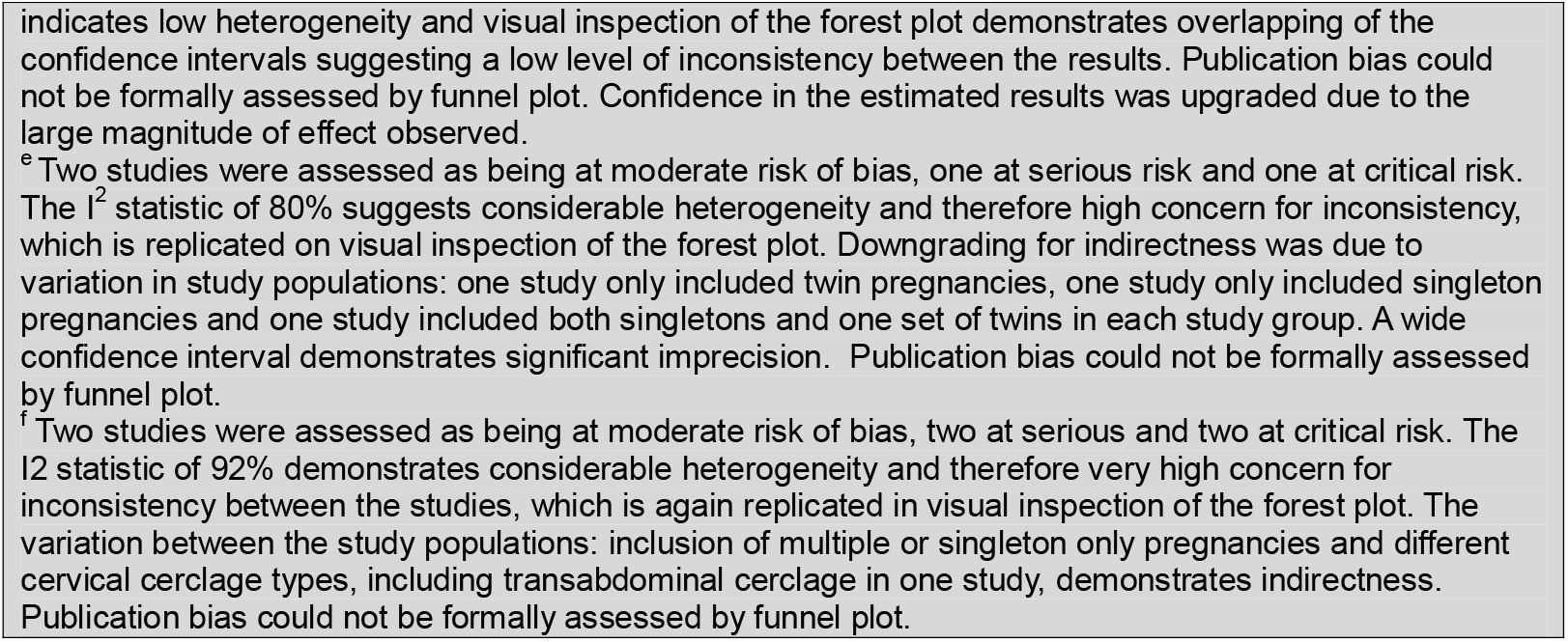
GRADE certainty of evidence assessment with explanations.

### 3.4 Data analysis

Meta-analyses were performed using RevMan software for each outcome that was reported by three or more studies: pregnancy loss as composite, preterm birth less than 37 weeks, preterm birth less than 34 weeks, miscarriage and previable neonatal death less than 24 weeks, birthweight in grams and gestational age at delivery in weeks. A summary of the meta-analyses are presented in table 4.

**Table 4:**
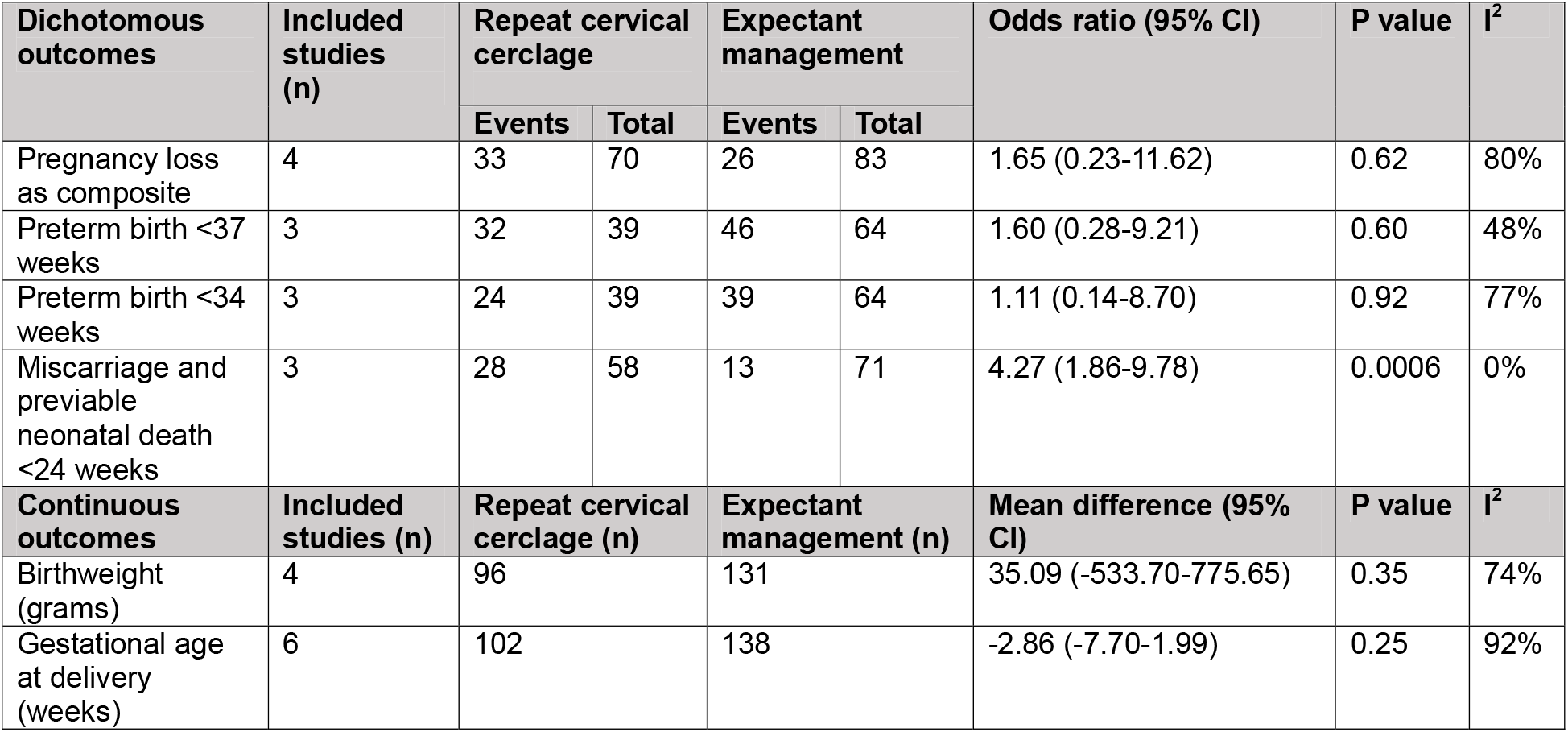
Summary of meta-analyses.

For pregnancy loss as composite, including four studies with 153 participants, (7, 16, 18, 19) the odds ratio (OR) was 1.65 with a 95% confidence interval (CI) of 0.23-11.62 and a p-value of 0.62, demonstrating no effect on pregnancy loss rate with or without repeat cervical cerclage when the primary cerclage fails (Fig 2).

**Figure 2:** Forest plot demonstrating pregnancy loss rate. Total number of participants used as denominator. Created using RevMan. Abbreviations: M-H = Mantel-Haenszel, CI = confidence interval

For preterm birth less than 37 weeks, including three studies with 103 participants, (17, 18, 20), the OR was 1.60 with a 95% CI of 0.28-9.21 and a p-value of 0.60, demonstrating no effect on preterm birth less than 37 weeks with or without repeat cervical cerclage (Fig 3).

**Figure 3:** Forest plot demonstrating preterm birth less than 37 weeks. Live birth rate used as denominator. Created using RevMan. Abbreviations: M-H = Mantel-Haenszel, CI = confidence interval

For preterm birth less than 34 weeks, including three studies with 103 participants, (17, 18, 20), the OR was 1.11 with a 95% CI of 0.14-8.70 and a p-value of 0.92, demonstrating no effect on preterm birth less than 34 weeks with or without repeat cervical cerclage (Fig 4).

**Figure 4:** Forest plot demonstrating preterm birth less than 34 weeks. Live birth rate used as denominator.

For miscarriage and previable neonatal death less than 24 weeks, including three studies with 129 participants, (7, 16, 18), the OR was 4.27 with a 95% CI of 1.86-9.78 and a P-value of 0.0006 (Fig 5). This shows a statistically significant association of repeat cerclage with increased odds of miscarriage and previable neonatal death less than 24 weeks.

**Figure 5:** Forest plot demonstrating miscarriage and previable neonatal death less than 24 weeks. Total number of participants used as denominator. Created using RevMan. Abbreviations: M-H = Mantel-Haenszel, CI = confidence interval

For birthweight, including four studies with 227 participants, (7, 17, 19, 20), the mean difference was 35.09g with a 95% CI of -533.70-603.89 and a p-value of 0.90, demonstrating no effect on birthweight with either repeat cervical cerclage or expectant management (Fig 6).

**Figure 6:** Forest plot demonstrating birthweight in grams. Total births used as denominator. Created using RevMan. Abbreviations: CI = confidence interval

For gestational age at delivery in weeks, including six studies with 240 participants, (7, 16-20), the mean difference was -2.86 with a 95% CI of -7.70-1.99 and a p-value of 0.25, demonstrating no effect on gestational age at delivery with either repeat cervical cerclage or expectant management.

**Figure 7:** Forest plot demonstrating gestational age at delivery in weeks. Total births used as denominator. Created using RevMan. Abbreviations: CI = confidence interval

### 3.5 Other secondary outcomes

Secondary outcomes were variably reported by our included studies, with particular paucity in secondary neonatal outcome data, and therefore we were unable to meta-analyse. Contag reported 8 out of 20 (40.0%) patients who received repeat cervical cerclage experienced PPROM, compared with 7 out of 31 (22.6%) from their expectant management group. (17) Tang conversely reported 3 out of 12 (25.0%) participants who received repeat cerclage experienced PPROM compared with 9 out of 24 (37.5%) who received expectant management. (20) Clarfield reported rates of placental infection being 92.9% in the repeat cerclage group and 66.7% in the expectant management group, although the clinical significance of this and how it was diagnosed was unclear. (7)

### 3.6 Subgroup analysis

A post-hoc subgroup analysis for pregnancy loss as composite was performed as during data extraction it was found that there was a clear divide between the definition used for primary cerclage failure; either a short cervix or prolapse of membranes beyond the level of the cerclage.

Only 4 studies were suitable for inclusion in the subgroup analysis and there was no evidence of a statistically significant difference between the subgroups (P=0.14). However, amongst those with a short cervix, repeat cerclage was associated with increased odds of pregnancy loss with an OR of 5.94 with a 95% CI of 1.25-28.27 and a p-value of 0.03. (Fig 8).

**Figure 8:** Forest plot demonstrating pregnancy loss rate according to subgroup of definition of cerclage failure. Total number of participants used as denominator. Created using RevMan. Abbreviations: M-H = Mantel-Haenszel, CI = confidence interval

## 4. Discussion

### 4.1 Summary of key findings

Repeat cervical cerclage insertion following failure of the primary cerclage has no association with pregnancy loss rate as composite, preterm birth less than 37 weeks and less than 34 weeks, birthweight and gestational age at delivery. The spread of effect, particularly for pregnancy loss rate and preterm birth, may suggest that worse outcomes are seen amongst those with repeat cerclage, compared to expectant management. For miscarriage and previable neonatal death less than 24 weeks, patients who receive repeat cervical cerclage may be four times more likely to suffer a pregnancy loss compared with those who receive expectant management (OR 4.27,95% CI 1.86-9.78). Those who received repeat cerclage for a short cervix on ultrasound follow-up had a significantly greater odds of suffering a pregnancy loss with repeat cervical cerclage compared to those receiving expectant management. There was no difference in pregnancy loss with or without repeat cerclage when failure was defined as prolapse of fetal membranes through the level of the cerclage. Overall, these results do not support the insertion of a repeat cervical cerclage when the primary cerclage fails.

### 4.2 Review of existing literature

Three additional papers were identified during screening that reported case series’ of repeat cervical cerclage following primary cerclage failure without a comparator group. Ru et al. reported the perinatal outcomes of 55 patients who underwent repeat cervical cerclage for: prolapse of membranes beyond the level of the cerclage but without cervical dilatation and cervical dilatation up to 6cm on physical examination. (21). Preterm birth less than 37 weeks was reported in 38 of 55 patients (69.1%) there was a 72.7% neonatal survival rate. Park et al. reported 35 women who underwent repeat cervical cerclage for prolapsed membranes on speculum examination following primary cerclage insertion. (22) Out of 35 patients, 8 had a miscarriage or previable neonatal death, 4 experienced what they describe as ‘perinatal death’, death less than 72 hours following birth and 5 had an early neonatal death, equating to a 48.6% rate of pregnancy loss as composite. Fox et al. reported 12 patients who underwent repeat cervical cerclage after primary cerclage. Their pregnancy loss rate (calculated as the inverse of live birth rate) was 1 out of 12 (8.4%) and 3 (25%) delivered prematurely before 37 weeks gestation.

These three studies do not offer comparison data and therefore, while descriptive of potential outcomes following repeat cerclage, they do not aid counselling or decision-making for whether repeat cerclage is beneficial compared to expectant management.

### 4.3 Strengths and limitations

This systematic review and meta-analysis was performed according to a prospectively published protocol, using robust methodology in line with the PRISMA 2020 guidelines. (8, 9) Comprehensive searches were carried out across the four main databases to ensure all possible studies eligible for inclusion were identified. To our knowledge there are no other published systematic reviews and meta-analyses of repeat cervical cerclage when the primary cerclage fails in asymptomatic patients.

The authors acknowledge that the quantity and quality of evidence used in this review is poor and therefore basing any recommendations based on this data alone is challenging. There are only retrospective cohort studies included each with varying degrees of bias; two at moderate risk, two at serious risk and two at critical risk. The certainty of evidence is low or very low, according to the GRADE assessments performed. It does, however, provide an important contribution to the body of evidence to guide and support clinical decision-making in this small subset of patients, where there are no pre-existing guidelines or robust data sets. Reporting of adverse maternal outcomes, such as PPROM, sepsis and chorioamnionitis was limited in the studies included in this review, and therefore there may be additional harm associated with repeat cervical cerclage that are not yet known. The sub-group analysis by definition of cerclage failure identified no significant difference between the groups which may reflect no true difference or underpowered to detect a difference given only two studies were able to be included in each subgroup. It is however noteworthy that in the women having repeat cerclage for a short cervix the risk of pregnancy loss was significantly increased compared to expectant management.

Given the retrospective nature of the studies, selection bias is likely to be significant, whereby patients with clinically more severe failure (e.g. greater dilatation of the cervix at the point of diagnosis of failure, or greatest risk according to their previous history) may have been selected to have a repeat cervical cerclage over expectant management. Clinicians may try everything possible to avoid pregnancy loss or extreme prematurity, particularly for those at greatest risk, and repeat cerclage may have been viewed as the last resort option. Although this is not demonstrated in the baseline population characteristics reported, it is clinically plausible and therefore may account for the trend seen towards worse outcomes for those who received repeat cervical cerclage compared to those who received expectant management.

### 4.4 Clinical application and recommendations

Routine ultrasound follow-up in the second trimester after cervical cerclage insertion has become more routine practice in UK Preterm Prevention Clinics in secondary and tertiary care. In terms of national and international guidance for ultrasound follow-up, the Society for Maternal-Fetal Medicine (SMFM) do not recommend routine ultrasound surveillance after cerclage insertion, as there is no evidence that repeat cervical cerclage improves outcomes. (23) The Royal College of Obstetricians (RCOG) similarly advise that routine ultrasound surveillance after history-indicated cervical cerclage is not recommended, although it can be offered to selected individuals where appropriate. (6)

As this review shows, inserting a repeat cervical cerclage may not be the most appropriate management plan following primary cerclage failure. However, timely identification of cerclage failure provides the opportunity for risk assessment and antenatal preparation. Particularly considering perinatal optimisation strategies with antenatal corticosteroids or in utero transfer to a centre with appropriate neonatal expertise, as per the British Association of Perinatal Medicine (BAPM) framework. (24) Identification of cerclage failure, particularly at gestations on the cusp of viability can guide clinicians to consider antenatal hospital admission, perform fetal growth assessment and provide the opportunity for neonatal counselling for parents. Without early identification of cerclage failure through routine ultrasound follow-up there is the risk that these patients attend symptomatic of preterm labour requiring emergency removal of their cerclage. They may also miss the window for optimally timed antenatal corticosteroids and in utero transfer to an appropriate neonatal intensive care centre.

### 4.5 Further research

This review was limited by the retrospective nature of the studies included. Future prospective cohort studies would provide much needed robust data to support our findings. An agreed definition for cerclage failure, or clear reporting of the indication for repeat cerclage is important for future meta-analysis. Although a randomized controlled trial would likely not be feasible due to the rare incidence of this clinical situation and therefore recruitment would be extremely challenging. In the UK there are several centres providing tertiary level Preterm Birth Prevention services, who comprise the majority of cerclage providers, and would be most likely to encounter this clinical situation. We aim to design a prospective cohort study in collaboration with these centres to examine outcomes from patients with cerclage failure, with or without repeat cervical cerclage insertion.

### 4.6 Conclusions

In conclusion, repeat cervical cerclage insertion when the primary cerclage fails, has no association with rates of pregnancy loss, preterm birth less than 37 weeks, preterm birth less than 34 weeks, birthweight or gestational age at delivery. There is a trend towards worse outcomes with repeat cerclage compared to expectant management, however it is unclear if the underlying risk of preterm birth at the time of primary cerclage failure differed between groups. The quantity and quality of the evidence base is poor and therefore patients should be informed of this during counselling. Prospective research studies in this area are needed to form more robust conclusions for optimal management following primary cerclage failure.

## Data Availability

All relevant data are within the manuscript and its Supporting Information files

## Acknowledgements

The authors would like to thank Derick Yates, Evidence Specialist at Birmingham Women’s and Children’s NHS Foundation Trust Library and Knowledge Services for his assistance with the search strategy and database searches. The authors would also like to thank the team at Birmingham Women’s and Children’s NHS Foundation Trust for their help retrieving full text articles.

