## Appendix 1 for "Should a repeat cervical cerclage be inserted when the primary cerclage fails, to prevent pregnancy loss and preterm birth? A systematic review and meta-analysis"

**Appendix 2: Secondary outcomes in full**

Maternal:

Gestational age at delivery including <28, <32, <34 and <37 weeks in live births ≥24 weeks gestation, time from conception to onset of spontaneous vaginal delivery, cerclage to delivery interval, mode of initiation of labour (spontaneous or induced), mode of delivery (vaginal, operative vaginal, caesarean section), vaginal bleeding (at placement of cerclage), antenatal steroid use, preterm pre-labour rupture of membranes (PPROM) and chorioamnionitis. Further secondary outcomes include, maternal mortality, maternal sepsis, maternal pyrexia (intrapartum and postnatal), maternal high dependency unit (HDU), or intensive treatment unit (ITU) admission (predelivery and postdelivery), and complications of cerclage placement: cervical laceration/tears (at placement and at delivery), bladder injury, iatrogenic rupture of membranes, insertion failure, repeat insertion, need of anaesthetic for removal, and difficult to remove sutures.

Neonatal:

Late neonatal death (>7 days but <28 days of life), neonatal death (early and late neonatal death inclusive), infant death (>28 days but <1 year of life), birthweight (in live births ≥24 weeks gestation), resuscitation at birth/additional care required (admission to special care baby unit (SCBU), neonatal intensive care unit (NICU), high dependency unit, or transitional care) and length of stay in additional care. Further outcomes: neonatal sepsis clinically diagnosed/proven (in live births ≥24 weeks gestation), gastrointestinal morbidity (including necrotising enterocolitis, in live births ≥24 weeks gestation), retinopathy of prematurity (in live births ≥24 weeks gestation), infant respiratory distress syndrome and respiratory support (ventilation/continuous positive airway pressure (CPAP)) including days on respiratory support (in live births ≥24 weeks gestation), supplemental oxygen requirements at 36 weeks postmenstrual age (in live births ≥24 weeks), neurodevelopmental morbidity including disabilities, cerebral palsy (in live births ≥24 weeks gestation), as well as long term outcomes after discharge.
