## Appendix 2 for "Should a repeat cervical cerclage be inserted when the primary cerclage fails, to prevent pregnancy loss and preterm birth? A systematic review and meta-analysis"

**Appendix 2: Example of full search strategy**

| 1. | exp Pregnancy/ |
| --- | --- |
| 2. | exp Pregnant Women/ |
| 3. | (pregnan* or gravid* or expectant mother or birthing person).ab,kw,ti. |
| 4. | 1 or 2 or 3 |
| 5. | ((cervix or cervical) adj (cerclage or stitch* or suture*)).ab,kw,ti. |
| 6. | (McDonald adj2 (stitch* or method or technique or suture* or cerclage)).ab,kw,ti. |
| 7. | (Shirodkar adj2 (stitch* or method or technique or suture* or cerclage)).ab,kw,ti. |
| 8. | 5 or 6 or 7 |
| 9. | exp Reoperation/ |
| 10. | (repeat or repetition or rescue or second or duplicat* or reinforc* or failure or failed).ab,kw,ti. |
| 11. | 9 or 10 |
| 12. | 4 and 8 and 11 |

Appendix 1: Example search strategy. “exp” denotes exploded MeSH headings. “ab.kw.ti” denotes searches within the abstract, keywords and title fields respectively.
