## Appendix 3 for "Should a repeat cervical cerclage be inserted when the primary cerclage fails, to prevent pregnancy loss and preterm birth? A systematic review and meta-analysis"

|  | Confounding | Selection of participants | Classification of interventions | Deviations from intended interventions | Missing data | Measurement of outcomes | Selection of the reported result | Overall risk of bias |
| --- | --- | --- | --- | --- | --- | --- | --- | --- |
| Baxter 2005 |  |  |  |  |  |  |  |  |
| Clarfield 2023 |  |  |  |  |  |  |  |  |
| Contag 2015 |  |  |  |  |  |  |  |  |
| Simcox 2012 |  |  |  |  |  |  |  |  |
| Song 2010 |  |  |  |  |  |  |  |  |
| Tang 2024 |  |  |  |  |  |  |  |  |

Grading of bias assessment for each study,

using the ROBINS-I tool.

Categories: low , moderate , serious ,

, serious

critical , no information
